# Prophylactic Antibiotics in Arteriovenous Fistulae: A systematic Review

**DOI:** 10.1101/2022.01.30.22270099

**Authors:** Ali Kordzadeh, Mekhola Hoff, Alan Askari, Ali D. Parsa

## Abstract

**Background:** The aim of this study is to establish and evaluate whether the use of prophylactic antibiotics in the creation of any autogenous arteriovenous fistula in hemodialysis patients is indicated, evidence-based and/or recommended.

**Methods:** A systematic review and meta-aggregation of the literature from 1966 to August 2016 in the English language in Medline, Scopus, Embase and Cochrane Library was conducted.

**Results:** The search produced a total of n=94 articles. Following the application of the recruitment criteria in accordance to PRISMA one (n=1) article was found eligible with a population of n=611 patients undergoing autogenous fistula formation. A total of n=136 patients received prophylactic antibiotics with no incidence of surgical site infection (SSI). The reported incidence of SSI in the group of patients (n=475) that did not receive prophylactic antibiotics was 0.2% (n=1). The quality of the article was assessed by the Oxford Critical Appraisal Skills Programme (CASP) and their recommendation for practice was evaluated through National Institute for Health and Care Excellence (NICE).

**Conclusion:** The first systematic review of the literature demonstrates that the current use of prophylactic antibiotics in the creation of any autogenous AVF is not evidence-based and further research in this area is highly advocated.

## Introduction

The autogenous arteriovenous fistula (AVF) is the preferred and gold standard modality of vascular access for patients with end stage renal disease requiring dialysis (1). The National Kidney Foundation Outcomes and Quality Initiative (NFK/DQQI) and Vascular Access Society (VAS) guidelines have set various parameters for the selection and creation of such fistulae for daily practice (1,2). However, no specific guideline and/or recommendation for the use of preoperative antibiotics in the formation of any autogenous AVF is available. The current recommendations refer to the use of antibiotics in arteriovenous grafts (AVG) and cryopreserved fistulae (3). It is believed that surgical site infection (SSI) in the creation of any autogenous AVF is rare but their occurrence could potentially be lethal in the presence of immunosuppression and significant comorbidities. Furthermore, such infections can result in distant infective embolization and on their detection immediate surgery is indicated. Currently, centers rely on anecdotal and expert opinion for the use of preoperative antibiotics in the creation of any autogenous AVF. In an era of emerging multidrug resistant microorganisms and financial strains on health care providers, a careful and evidence based practice could prove beneficial in daily practice. Therefore, the aim of this study is to establish and evaluate whether the use of prophylactic antibiotics in the creation of any autogenous arteriovenous fistula in hemodialysis patients is indicated, evidence based and/or recommended. To the best of our knowledge this is the first systematic review of the literature that has evaluated this topic.

## Materials and methods

### Literature search

An electronic search of Medline, Scopus, Embase and Cochrane Library and Complementary Medicine Database (AMED), Health Management Information Consortium (HMIC), British Nursing Index (BNI) from 1966 to August 2016 was carried out using the following MeSH terms: arteriovenous fistula; antibiotic prophylaxis; prophylactic antibiotic; preoperative antibiotic. Studies appearing to fulfil the eligibility criteria but insufficient information within the abstracts were also retrieved and examined in full. This search was limited to adult subjects and English language only.

### Selection Criteria

The following restrictions were applied and following studies were excluded: 1) Any studies that included any type of fistulae created with non-autogenous material 2) Animal studies 3) Experimental studies with/or no endpoint 4) Case reports 5) Conference proceedings and commentaries.

### Search Outcome

This systematic search produced a total of n=94 hits. All abstracts were retrieved and reviewed by two separate investigators (MH and AK). Studies appearing to fulfil the eligibility criteria but possessed insufficient information within the abstracts were also retrieved and examined in full. Data extraction was performed by two separate investigators (MH and AK). This was performed in accordance to Preferred Reporting Items for Systematic Reviews and Meta-analysis (PRISMA) (4) (Figure 1). This resulted in only one retrospective cohort article with a population of n=611 patients.

**Figure 1.**
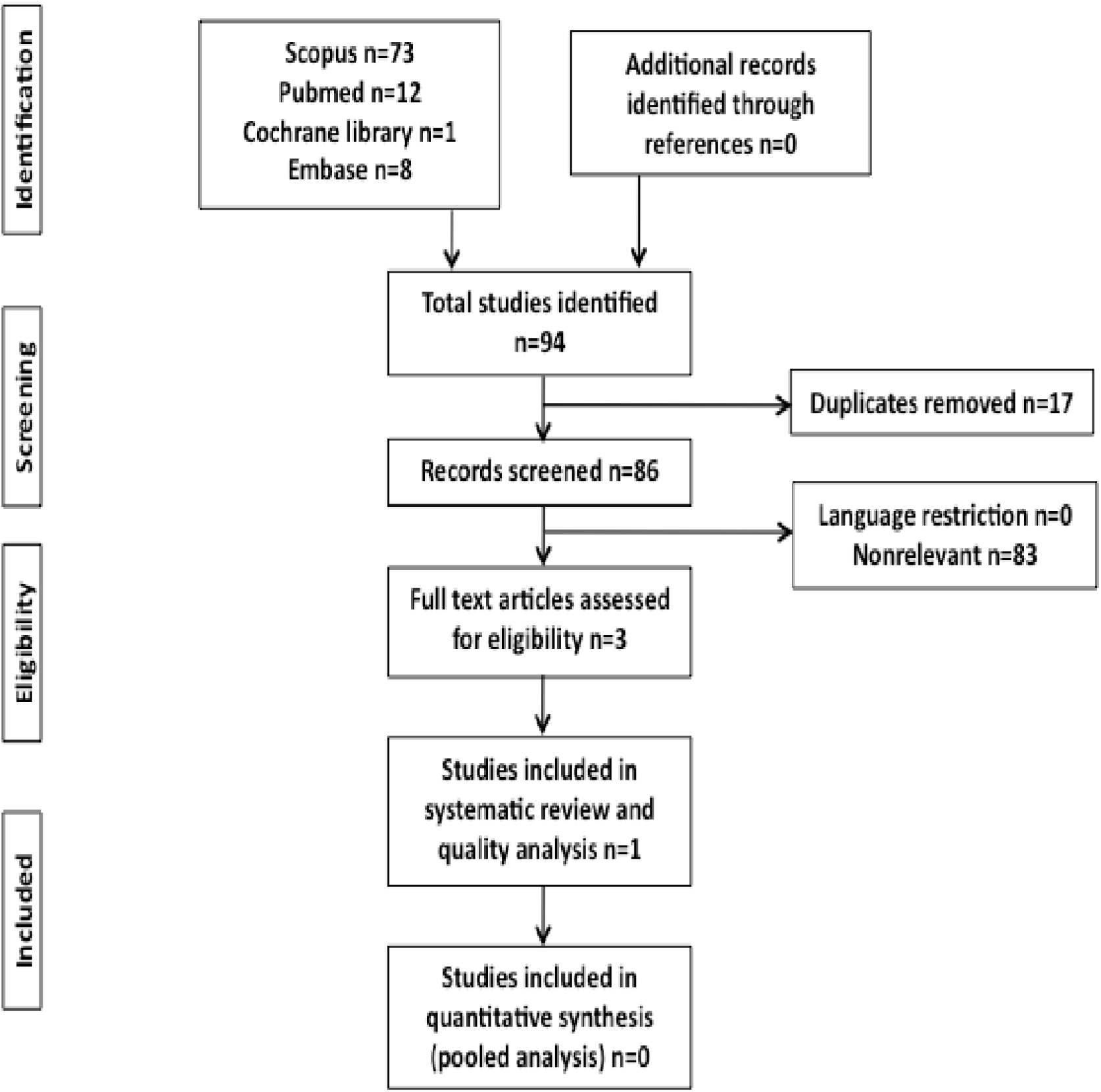
Flow chart for this systematic review (PRIMSA).

### Quality Assessment and Analysis

In order to achieve an evidence-based approach and reach an informative conclusion, the included article was assessed for its validity, bias, applicability and importance through the Critical Appraisal tool provided by the Oxford Critical Skills Programme (CASP) (5). Due to a lack of adequate articles a meaningful statistical analysis was not plausible. Therefore, the data was presented in a tabulated format and discussed in detail. The strength of evidence and their recommendations for practice was also assessed through National Institute for health and care Excellence (NICE) (6).

## Discussion

One article by Nagai et al. (7) was found eligible for this systematic review. This retrospective cohort study evaluated the role of prophylactic antibiotics to the incidence of surgical site infection (SSI) in n=651 patients post autogenous AVF, AVG and PD catheter placement. However, for the purpose of this systematic review the focus will be on autogenous AVFs. The aforementioned study, had three different phases. In phase one, a total of n=120 patients undergoing autogenous AVF were evaluated for SSI with preoperative single dose prophylactic antibiotic. The study end point and observation period was set from creation to 2 weeks postoperatively. During this period no SSI was identified in any of the individuals. In phase two of the study, a comparative study between two groups of patients undergoing autogenous AVF was conducted. In this phase, n=16 individuals received prophylactic antibiotics whereas n=22 patients did not receive any. The study end point remained similar to the first phase. Both groups exhibited similar age, (61 vs 64 years), gender predominance (female n=6 vs 7), serum albumin, and protein levels. The incidence of diabetes mellitus (DM) was also similar (DM n=7 vs n=11). The outcome revealed one case of possible SSI (only redness after 5 days) in the group that did not received any prophylactic antibiotics. In the third phase of the study, an observational study was performed on n=453 cases of autogenous AVF. The time to SSI was also set from creation to 2 weeks postoperatively. None of these individuals received any prophylactic antibiotics and no SSI was reported. Overall, n= 475 individuals did not receive any antibiotics and only one reported case of suspicious SSI was noted (n=1/475, 0.2%).

The study by Nagai et al. (7) could be subject to a degree of performance bias, as no information regarding the type of autogenous AVF, preparatory skin solution, duration of surgery and length of hospital stay was provided to the readers. However, given all patients were from a single institution, a logical assumption can be formulated that exposure of all individuals undergoing autogenous AVF in both groups of prophylactic and non-prophylactic group (n=136 vs n=475) were similar and/ or spread in terms of aforementioned factors. In addition, the number of recruited cases (power) becomes the strength of this article in excluding conclusion bias (type I and II). The investigators also paid significant attention to the maintenance and competent airflow and filtration system of their operating theaters. These factors remain vital in the era where high turnover and increased demand for productivity against fiscal years in national health providers are considered.

The incidence of infection in clean site surgery, like that of autogenous fistulae, is relatively low and the requirement of short and prolonged antibiotics is not defined and/or currently recommended. In our unit, we do not routinely administer prophylactic antibiotics for the creation of autogenous radiocephalic (RCAVF) and /or brachiocephalic fistulae (10). Our experience shows SSI following such procedures are negligible and limited to severely immunocompromised individuals and prophylactic antibiotics are not clinically and/or financially justified. However, in scenarios where surgery like that of basilic vein mobilization and/or complex autogenous loop formation could be prolonged and/or complicated, the use of antibiotics (prophylactic) remains open to debate. In such circumstances, there are two valid arguments. Firstly, the formation of complex access indicates the lack of other options (RCAVF, BCAVF) and the importance of the fistulae, albeit minimization of possible complications like that of infection, might be advised. Therefore, single dose prophylactic antibiotic might be indicated and if not this should be balanced against the likely risk of developing a post-operative infection, which itself can require prolonged antibiotic with its concomitant side effects, allergic reactions, prolonged hospital stay, morbidity, mortality and cost implications to the health care provider. Lastly, some surgeons believe attention and meticulous preparation in combination to shortened operative period performed by an experienced surgeon could avoid postoperative complications such as SSIs. However, in complex cases where the axilla or axillary fold is exposed, the incidence of infection could escalate.

In order to avoid SSIs, consideration of other factors must also be taken into account. For instance, patients could be screened for methicillin resistant staphylococcus aureus (MRSA) prior to surgery, and eradication programme could be started early. Another factor is related to the type of preparatory skin solution. This varies from center to center but recent Cochrane database suggests superior outcomes with chlorhexidine alcohol for skin preparation than povidone iodine (8,9). Significant attention should be paid to patients that are immunocompromised or might have had previous records of infection and/or suffer from significant comorbidities.

## Conclusion

Finally, the level of evidence (Level 2, Grade c) derived from the current systematic review is not sufficient to make a solid recommendation for daily practice.

However, it appears that the incidence of SSI without prophylactic antibiotics in autogenous AVF formation is only limited to 0.2% according to this systematic review. However, their routine use in more complex cases remains open to debate and dependent on each unit’s expertise and experience (10).

### Recommendation for Practice

- The reported incidence of SSI for non complex autogenous AVF is limited to 0.2%.
- The routine use of prophylactic antibiotics in non complex autogenous AVF is not supported by literature.

### Recommendation for Research

- Randomised controlled trial in this field is highly advocated.
- Further research is required to investigate the type of autogenous AVF, preparatory skin solution, duration of surgery and length of hospital stay in combination with patient demographics, comorbidities and expertise.

## Data Availability

This is a review of the present evidence in the literature and all data are available on line.

## Disclosure

Financial Support: The authors have no financial disclosure to make.

Conflict of Interest: The authors have no conflict of interest.

Ethical Approval : Not applicable as this is review of evdience.

---

**Table 1.**
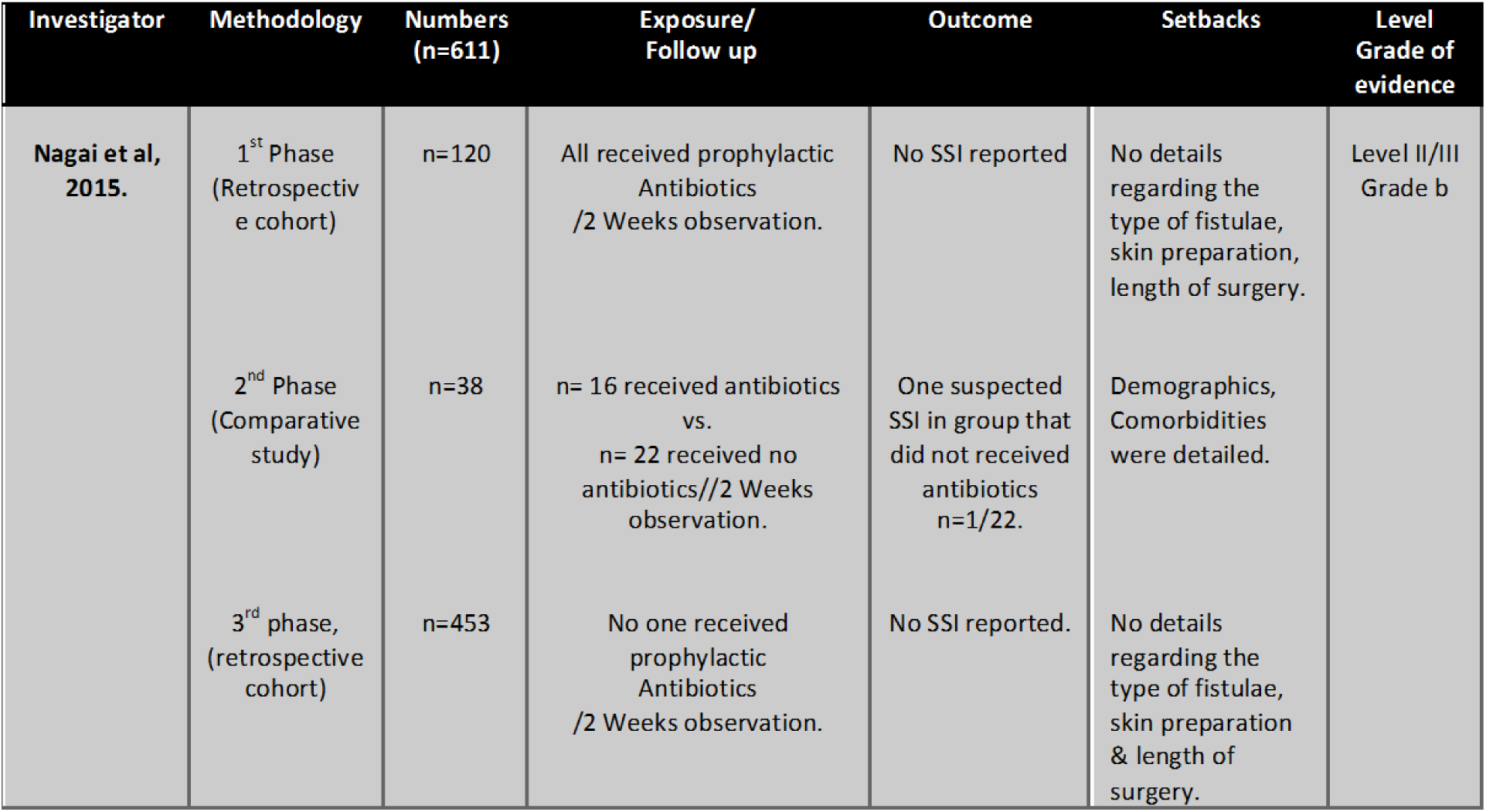
Summary of the systematic review.

